# A simplified SARS-CoV-2 pseudovirus neutralization assay

**DOI:** 10.1101/2021.03.12.21253435

**Authors:** Gaetano Donofrio, Valentina Franceschi, Francesca Macchi, Luca Russo, Anna Rocci, Valentina Marchica, Federica Costa, Nicola Giuliani, Carlo Ferrari, Gabriele Missale

## Abstract

COVID-19 is an ongoing pandemic caused by the highly infectious coronavirus SARS-CoV-2 that is engaging worldwide scientific research to find a timely and effective eradication strategy. Great efforts have been put into anti-COVID-19 vaccine generation in an effort to protect the world population and block SARS-CoV-2 spread. To validate the protective efficacy of the vaccination campaign and effectively control the pandemy, it is necessary to quantify the neutralizing antibodies induction by vaccination, since they have been established to be a correlate of protection. In this work a SARS-CoV-2 pseudovirus neutralization assay, based on a replication incompetent lentivirus expressing an adapted form of CoV-2 S protein and an ACE2/TMPRSS2 stably expressing cell line, have been minimized in term of protocol steps without loss of accuracy. The goal of the present simplified neutralization system is to improve SARS-CoV-2 vaccination campaign by means of an easy and accessible approach to be performed in any medical laboratory, maintaining the sensitivity and quantitative reliability of classical serum neutralization assays. Further this assay can be easily adapted to different coronaviruses variants by simply modifying the pseudotyping vector.

## Introduction

Coronaviruses, belonging to the family Coronaviridae in the order Nidovirales, are positive-strand RNA viruses with a genome length comprised between 26 and 32 Kbp. Several mammalian and avian species can be infected by coronavirus and most of the time causing respiratory and/or intestinal disease [1]. Human coronaviruses (HCoVs), such as HCoV-229E, HCoV-OC43, HCoV-NL63, and HKU1, have long been recognized as major causes of the common cold and are endemic in the human population. Two recent HCoVs, severe acute respiratory syndrome coronavirus (SARS-CoV) and Middle East respiratory syndrome coronavirus (MERS-CoV), emerged in 2002 and 2012, respectively, causing life-threatening disease in humans. A previously unknown coronavirus, named SARS-CoV-2 (CoV-2), was discovered in December 2019 in Wuhan, China, responsible for a pandemic infection, known as coronavirus disease 19 (COVID-19) and causing a large number of deaths people worldwide [1]. Despite a huge research effort has been made, COVID-19 remains a complex disease showing pathogenetic mechanisms and clinical heterogeneous features difficult to understand. A variety of approaches have been employed to develop prophylactic and therapeutic measures, including whole inactivated vaccines, subunit vaccines, RNA-based vaccines, viral vectored vaccines [2,3], monoclonal neutralizing antibodies and fusion inhibitors, most of which were designed to target the CoV-2 Spike glycoprotein (S) [4,5]. CoV-2 S, forming homotrimers structures on viral surface, mediates virus entry to the host cell. S mature structure is formed by S1 and S2 functional subunits: S1 interacts with the Angiotensin Converting Enzyme 2 (ACE2) cellular receptor, while S2 mediates the viral envelope fusion with the host cell membrane [6-8]. During S1-ACE2 interaction, the host cell surface Transmembrane Serine Protease 2 (TMPRSS2), located next to ACE2 receptor, cleaves the S2 subunit at the S2’ amino terminal portion (815↓816aa; SKR↓SFI), inducing the fusion peptide hydrophobic domains exposure and the subsequent viral envelope fusion with the host cell membrane [6-8]. Due to its high infectivity and pathogenicity, CoV-2 needs to be handled in biosafety level 3 (BSL-3) specific facilities (https://www.cdc.gov/coronavirus/2019-ncov/lab/lab-biosafety-guidelines.html) [9], which limits the development of anti-viral measures as well as basic and applied studies on the interaction between host cells and CoV2 and viral attachment and entry mediated by the S protein. To avoid dealing with infectious CoV2, several safe, biosafety level 2 (BSL2) pseudovirus-based systems have been developed mainly based on vescicular stomatitis virus (VSV) [10] or retrovirus (RV) [11,12] vector pseudotyped with CoV-2 S. Although both of them have been shown to be sensitive and reliable, they suffer to be farraginous, time consuming and expensive in procedural terms. In the present work, a 4 steps simplified procedure of CoV-2 pseudovirus neutralization assay was established.

## Material and Methods

### Plasmids

ACE2-IRES-TMPRSS2-IRES-Puromycin tricistronic ORF was chemically synthetized and integrated into a lentiviral transfer vector to get pEF1α-ACE2/TMPRRS2/Puro (Supplementary Figure 1 for details and full sequence). Similarly, S-ΔRS-HA ORF (Supplementary Figure 2) and biscistronic turboGFP-IRES-Luc2 ORF were chemically synthetized and integrated into a lentiviral transfer vector to get pLV-CMV-(S-ΔRS-HA)-IRES-Puro-WPRE (Supplementary Figure 3) and pLV-EF1α-(turboGFP-IRES-Luc2)-WPRE respectively. p8.74 packaging, pREV, pMD2 pseudotyping and pEGFP-C1 vectors were obtained from Addgene (https://www.addgene.org/).

### Cells

Human Embryo Kidney (HEK) 293T (ATCC: CRL-11268) cells were cultured in Eagle’s Minimal Essential Medium (EMEM, Gibco; Thermo Fisher Scientific, Carlsbad, CA, USA) containing 1mM of Sodium Pyruvate (Gibco), 2 mM of L-glutamine (Gibco), 100 IU/mL of penicillin (Gibco), 100μg/mL of streptomycin (Sigma-Aldrich, Milano, Italy), and 0.25μg/mL of amphotericin B (Gibco); called complete EMEM] supplemented with 10% Fetal Bovine Serum (FBS, Gibco), and were incubated at 37 °C and 5% CO_2_ in a humidified incubator.

Stably transfected HEK/S-ΔRS-HA/Puro (HEK/S) and HEK/ACE2/TMPRRS2/Puro cells were obtained transfecting cells with pLV-CMV-(S-ΔRS-HA)-IRES-Puro-WPRE or pEF1α-ACE2/TMPRRS2/Puro vectors, respectively. Briefly, sub-confluent HEK 293T cells were detached from a T75cm^2^ flask, counted and electroporated with 20 µg of pLV-CMV-(S-ΔRS-HA)-IRES-Puro-WPRE or pEF1α-ACE2/TMPRRS2/Puro vectors in 600 µL of DMEM high glucose (Euroclone) without serum at 186V, 1500µF in Gene Pulser XCell (Biorad, Milano, Italy).

Electroporated cells were then transferred to new 25 cm^2^ flasks and fed with complete EMEM with 10% FBS. Twenty-four hours after the transfection, the medium was changed with fresh complete EMEM with 10% FBS complemented with 2 µg/ml of puromycin (Millipore Merck Life Science, Milano, Italy). Cells were kept in culture until resistant colonies appeared. Cells were split for more than 40 passages and tested for S or ACE2 expression.

### Transient Transfection and syncytia formation

HEK 293T cells were transiently co-transfected with pLV-CMV-(S-ΔRS-HA)-IRES-Puro-WPRE, pEF1α-ACE2/TMPRRS2/Puro and pEGFP-C1 (Clontech) vectors, with same molar ration (1:1:1), using polyethylenimine (PEI) transfection reagent (Polysciences, Inc. Warrington, Pennsylvania, USA). Briefly, cells were seeded at 5×10^5^cells/well in 6-well plates and incubated overnight at 37 °C/5% CO2. Cells were then incubated for 6 hours with a transfection mix (1 ml) containing 3μg of plasmids DNA and PEI (ratio 1:2.5 DNA-PEI) in complete DMEM (Dulbecco’s Modified Essential Medium (DMEM) high glucose (Euroclone) completed with 50 μg/ml of Gentamicine (Merk)) without serum. After incubation, the transfection mix was replaced by fresh complete EMEM and incubated for 24 hours at 37°C, 5% CO2. HEK/ACE2/TMPRRS2/Puro cells were transiently co-transfected with pLV-CMV-(S-ΔRS-HA)-IRES-Puro-WPRE and pEGFP-C1 as before. Alternatively, HEK/ACE2/TMPRRS2/Puro cells were also co-cultivated (1:2 ratio) with HEK 293T cells transiently transfected with pLV-CMV-(S-ΔRS-HA)-IRES-Puro-WPRE and pEGFP-C1. Twenty-four hours post co-cultivation, syncytia were observed by inverted fluorescence microscope (Zeiss-Axiovert-S100) and pictures were acquired by digital camera (Zeiss-Axiocam-MRC). HEK/S-ΔRS-HA/Puro and HEK/ACE2/TMPRRS2/Puro cells were also co-cultivated as before to generate syncytia.

### Western Immunoblotting

Protein cell extracts were obtained from pLV-CMV-(S-ΔRS-HA)-IRES-Puro-WPRE, and pEGFP-C1 transfected HEK293T cells by adding 100μL of cell extraction buffer (50 mM Tris-HCl,150 mM NaCl, and 1% NP-40; pH 8). After BCA total protein quantification (Pierce™ BCA Protein Assay kit, Thermo Fisher Scientific), cell extracts containing various amount of total protein were electrophoresed through 10% SDS-PAGE. Proteins were then transferred to a nylon membrane by electroblotting, and the membrane was incubated with mouse monoclonal antibody anti-HA tag (G036, Abm Inc., Vancouver, Canada) diluted 1:10,000. After washing, the membrane was incubated with a rabbit anti-mouse IgG secondary antibody labelled with horseradish peroxidase, diluted 1:15,000 (A9044, Sigma-Aldrich) and visualized by enhanced chemiluminescence (Clarity Max Western ECL substrate, Bio-Rad, Hercules, California, USA).

### Hematoxylin and Hematoxylin and Eosin staining

Flasks of cells containing syncytia were fixed with 4% of paraformaldehyde in PBS and stained with Hematoxylin and Eosin standard method.

### Pseudovirions reconstitution

For reconstituting CoV-2 S pseudovirus, HEK 293T cells were transfected, in T175 cm^2^ flasks, with 25 μg of pLV-EF1α-(turboGFP-Luc2)-WPRE transfer vector, 15 μg of p8.74 packaging vector, 13 μg of pLV-CMV-(S-ΔRS-HA)-IRES-Puro-WPRE pseudotyping vector (although this is a transfer vector, it can work as a pseudotyping vector too) and 5μg of pREV (58 μg of total DNA), were diluted in 3 mL of complete DMEM (Euroclone) without serum and 145 μL of PEI (Polysciences, Inc.) (1 mg/mL in PBS) (ratio 1:2.5 DNA-PEI). After at least 15 minuts incubation at room temperature, 4x volumes of complete DMEM without serum were added and the transfection solution and transferred to the cell monolayer. After 6 hours of incubation at 37°C/5% CO_2_, in a humidified incubator, the transfection mixture was replaced with 25 mL of fresh complete EMEM supplemented with 10% FBS and incubated for 48 hours at 37 °C/5% CO_2_. The flask was then frozen-thawed at −80°C, Transfected Cells Supernatant (TCS) containing S Pseudovirus was clarified via centrifugation at 3,500 rpm for 5 minutes at 4°C, filtered through a 0.45 μm filter (Millipore), aliquoted, tittered by limited dilution and stored at −80 °C.

ACE2/TMPRSS2 pseudovirus was prepared as described above by simply substituting pLV-CMV-(S-ΔRS-HA)-IRES-Puro-WPRE with pEF1α-ACE2/TMPRRS2/Puro, again, although this is a transfer vector, it can work as a pseudotyping vector too.

### Serum samples

Sera were collaboratively provided by the Unit of Infectious Diseases and Hepatology, University Hospital of Parma. The study was approved by the local ethical committee (Comitato Etico Area Vasta Emilia Nord (AVEN), Italy). All participants gave written informed consent to participate in the study.

### Seroneutralization assay

25µL of complete EMEM with 10% of FBS were added to each well of a 96 wells opaque wall, clear flat bottom white microplate (Greiner bio one) and 25 µL of each serum were added to the first line of wells (**Supplementary Figure 5**). 25µL of S pseudovirus preparation (described in Pseudovirions reconstitution section above) diluted in complete EMEM with 10% of FBS (corresponding to ∼10^4^ Relative Luciferase Units (RLUs); ∼3-5µL of the initial preparation) were added to each well and left to incubate at room temperature for 1.5 hour. Final volume for each well went up to 50µL, therefore the sera dilution was doubled, 1:4-1:8-1:16-1:32-1:64-1:128-1:256-1:512. Next, 50µL of complete EMEM with 10% FBS, containing 10^4^ HEK/ACE2/TMPRRS2/Puro cells wear added to each well and left for 60 hours at 37 °C/5% CO_2._

Plates were read by adding 25µL of complete EMEM containing Luciferine to each well just before the reading of the microplate to the Luminometer (Victor, Perkin Elmer). Since the plate has a transparent bottom, a dark film layer should be applied for reading. A negative control was established without serum, which was substituted with complete medium. The RLUs were compared and normalized to those derived from wells where pseudovirus was added in the absence of sera (100%). Neutralization Titer 50 (NT50) was expressed as the maximal dilution of the sera where the reduction of the signal is ≥50%. Note of worthy, the titer has to be multiplied per 40 because the initial volume of the sera tested is 0,025mL and it has to be normalized to 1mL.

## Results and discussion

### Generation of a sensitive target cell line simultaneously expressing ACE2 and TMPRRS2

Since ACE2 and TMPRSS2 are the main restriction factors for SARS-CoV-2 (CoV-2) attachment and penetration into the host cells [7], a cell line simultaneously expressing ACE2 and TMPRSS2 was generated. For this purpose, a tricistronic expression cassette was constructed. Human ACE2 ORF (GeneBank accession number: NM_021804.3), provided of a strong Kozak’s sequence to the 5’ end, human TMPRSS2 ORF (GeneBank accession number: NM_001135099.1) and the Puromycin drug resistance gene ORF were in-tandem positioned and linked by two Internal Ribosomal Entry Sites (IRES) (ACE2-IRES-TMPRSS2-IRES-Puromycin). This large tricistronic ORF was chemically synthetized and sub-cloned downstream to the human Elongation Factor 1 alpha promoter (EF1α), upstream to the SV40 polyA signal/site in a pUC plasmid backbone. Thus, pEF1α-ACE2/TMPRRS2/Puro was generated (Supplementary Figure 1). pEF1α-ACE2/TMPRRS2/Puro, was electroporated in HEK293T cells and stably transfected cells were obtained by selection with puromycin. Since puromycin drug resistance gene ORF is the last to be translated from the mRNA transcribed from pEF1α-ACE2/TMPRRS2/Puro, the selection of puromycin selected resistant cells (HEK/ACE2/TMPRRS2/Puro) indeed guaranteed the expression of ACE2 and TMPRSS2. HEK/ACE2/TMPRRS2/Puro cells were kept, so far, for over thirty passages without loss of functionality/expression. Noteworthy, HEK/ACE2/TMPRRS2/Puro cells do not need to be constantly maintained in culture, under puromycin selection: a large batch of them can be grown, aliquoted at a suitable concentration, stored at −80 C° or in liquid nitrogen and successively used, when needed, by simply thawing an aliquot of them, without any loss of functionality.

### Expression of SARS-CoV-2 Spike glycoprotein in mammalian cells

CoV-2 S is a key factor for SARS-CoV-2 target cell infection [6-8]. Antibodies from COVID-19 convalescent patient sera can block *in vitro* and *in vivo* CoV-2 infectivity, thus being exploitable for CoV-2 infection diagnosis and for the evaluation of CoV-2 vaccination efficacy (the titer of neutralizing antibody present in a serum is a correlate of protection), as well as for preliminary test molecules able to interfere with S, ACE2 and TMPRRS2 interaction. Before attempting the generation of a Lenti-based pseudovirus displaying CoV-2 S on its surface, it was indispensable to generate an appropriate expression cassette capable to drive an efficient expression of S, accommodated on the pseudovirus envelope surface. The cytoplasmic tails of some CoV S proteins contain an Endoplasmic Reticulum Retrieval Signal (ERRS) that can retrieve S proteins from the Golgi to the Endoplasmic Reticulum (ER). This process is thought to accumulate S proteins at the CoV budding site, the ER-Golgi Intermediate Compartment (ERGIC), and to facilitate S protein incorporation into virions [13,14]. In contrast to Coronaviruses, Lentiviruses assemble and buds at the cell surface [15], therefore truncation of the S-protein cytoplasmic tail may increase cell surface levels and/or enable incorporation by alleviating structural incompatibility of the S-protein cytoplasmic tail and lentivirus proteins. CoV-2 S ORF sequence (https://www.ncbi.nlm.nih.gov/protein/1791269090) was deprived of its last 57 bp, coding for a predicted ERRS (KFDEDDSEPVLKGVKLHYT) [13] and substituted with the Hemoagglutinin (HA) tag. The so designed ORF (S-ΔRS-HA) was human codon usage adapted, with the use of the Jcat codon adaptation tool (http://www.jcat.de/), to change nucleotide codon composition to a composition based on human genome codon usage. The degeneracy of the genetic code leads to a situation whereby most of the amino acids can be encoded by two to six synonymous codons. The synonymous codons are not equally utilized to encode the amino acids, thus resulting in phenomenon of codon usage bias. Since codon usage bias has been shown to correlate with gene expression level, it has been proposed as an important design parameter for enhancing recombinant protein production in heterologous host expression [16]. The GC content of S-ΔRS-HA is 37% and adaptation to the human genome codon usage shifted the GC content from 37% up to 63% (Supplementary Fig. 2). Although we did not compare the adapted S-ΔRS-HA ORF with the un-adapted one in terms of expression efficiency, previous studies have shown that GC-rich genes in mammalian cells can be expressed 100-fold more efficiently than their GC-poor counterpart due to increased steady-state mRNA levels [17]. S-ΔRS-HA was chemically synthetized and integrated in a transfer vector under the transcriptional guide of the Immediate Early gene promoter of human Cytomegalovirus (CMV) and followed by an IRES, the puromycin drug resistance gene and the Woodchuck Hepatitis Virus (WHP) Posttranscriptional Regulatory Element (WPRE) (pLV-CMV-(S-ΔRS-HA)-IRES-Puro-WPRE (Fig. 1A and Supplementary Fig. 3 for complete sequence).

**Fig. 1.**
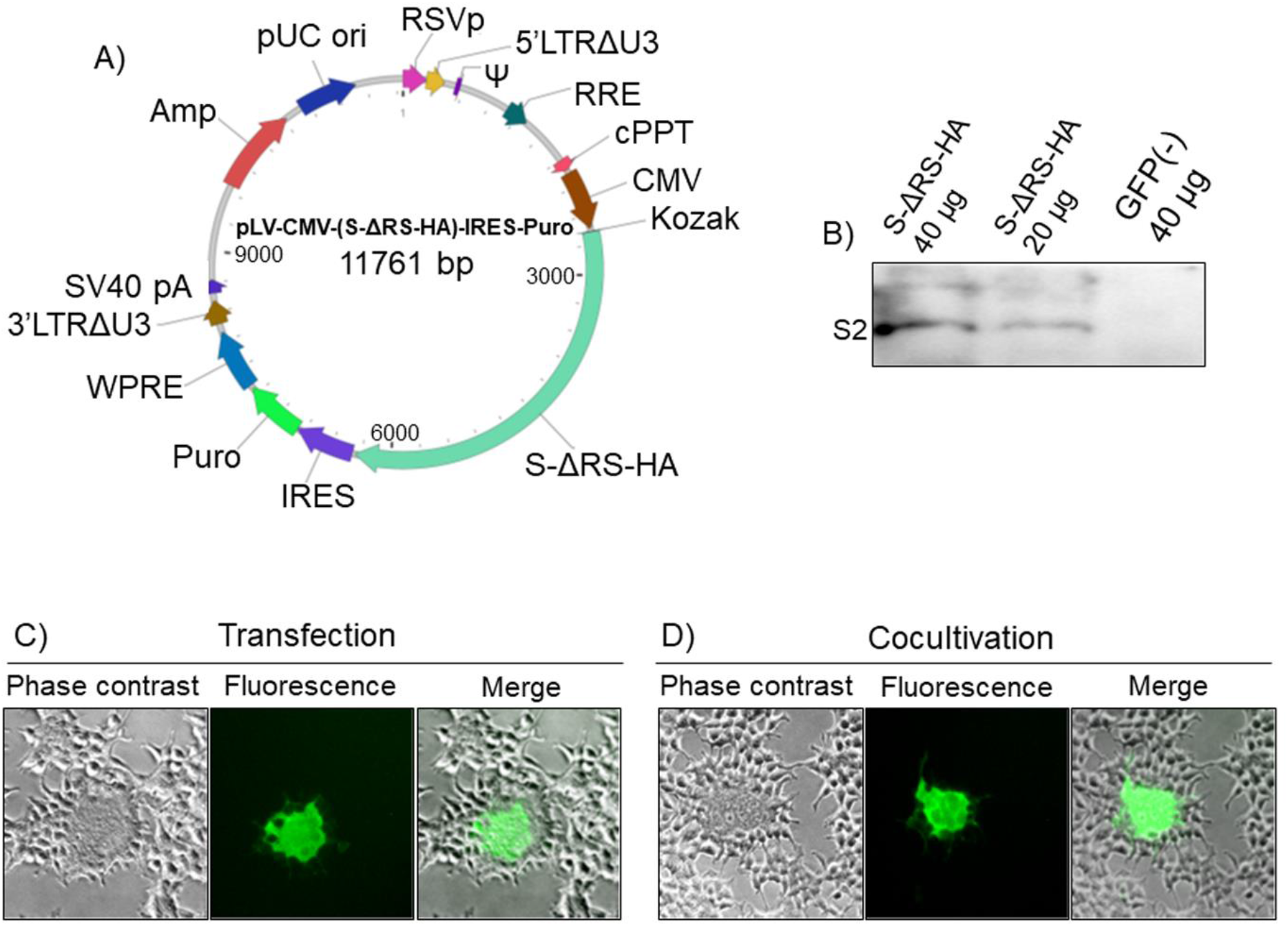
CoV-2 S expression in HEK293T cells. A) Diagram not on scale of pLV-CMV-(S-ΔRS-HA)-IRES-Puro-WPRE transfer vector, where functional elements of the construct are indicated and highlighted with different colours. RSVp (Rous sarcoma virus enhancer/promoter), 5’LTRΔU3 (Truncated HIV-1 5’ Long terminal repeat), Ψ (HIV-1 packaging signal, RRE (HIV-1 REV response element), cPPT (Central polypurine tract), CMV (Human cytomegalovirus immediate early enhancer/promoter), Kozak (Kozak’s translation initiation sequence), S-ΔRS-HA (Human codon usage adapted, HA tagged and retention signal delete CoV-2 S ORF), IRES (Encephalomyocarditis virus internal ribosomal entry site), Puro (Puromycin resistant gene ORF), WPRE (Woodchuck hepatitis virus posttranslational regulatory element), 3’LTRΔU3 (Truncated HIV-1 3’ long terminal repeat; self-inactivation), SV40 early pA (Simian virus 40 early polyadenylation signal), Amp (Ampicillin resistance gene, comprising the promoter and ORF) and pUC ori (High copy number pUC origin of replication). B) Western immunoblotting of pLV-CMV-(S-ΔRS-HA)-IRES-Puro-WPRE transfected HEK293T cells protein extracts (S-ΔRS-HA; 40 and 20 µg) and pEGFP-C1 transfected HEK293T cells protein extract (GFP; 40 µg) employed as a negative control (-). C) Representative microscopic image (Phase contrast, Fluorescence and Merged fields; 10X) of syncytia, generated by co-transfection of HEK/ACE2/TMPRRS2/Puro cells with pLV-CMV-(S-ΔRS-HA)-IRES-Puro-WPRE construct and PEGFP-C1. D) Representative microscopic image (Phase contrast, Fluorescence and Merged fields; 10X) of syncytia, generated by co-cultivation of HEK/ACE2/TMPRRS2/Puro cells with pLV-CMV-(S-ΔRS-HA)-IRES-Puro-WPRE and pEGFP-C1 co-transiently transfected HEK293T cells.

pLV-CMV-(S-ΔRS-HA)-IRES-Puro-WPRE transfected cells successfully expressed S-ΔRS-HA protein as shown by western immunoblotting (Fig. 1B). When HEK/ACE2/TMPRRS2/Puro where transiently cotransfected with pLV-CMV-(S-ΔRS-HA)-IRES-Puro-WPRE and pEGFP-C1, a construct delivering Enhanced Green Fluorescent Protein (EGFP), large syncytia were observed (Fig. 1C). This syncytiogenic effect was attributable to the interaction between S, ACE2 and TMPRRS2, which recapitulated the bonafide fusogenic activity of S when is attached to ACE2, proteolytically activated by TMPRSS2 and regulated by Interferon-Induced Transmembrane (IFITMs) proteins [18,19]. An identical result was obtained when HEK/ACE2/TMPRRS2/Puro cells were co-cultivated with pLV-CMV-(S-ΔRS-HA)-IRES-Puro-WPRE and pEGFP-C1 co-transiently transfected HEK293T cells (Fig. 1D). Further, syncytia could be reduced or abrogated if pLV-CMV-(S-ΔRS-HA)-IRES-Puro-WPRE transfected HEK/ACE2/TMPRRS2/Puro cells were treated with human COVID-19 convalescent sera containing anti-S antibodies or TMPRSS2 inhibitors (data not shown).

### Generation of a lenti-based pseudovirus

Relying on the promising results obtained with pLV-CMV-(S-ΔRS-HA)-IRES-Puro-WPRE construct and HEK/ACE2/TMPRRS2/Puro cell line, the reconstitution of a second generation replicating incompetent lentivirus, pseudotyped with CoV-2 S was attempted. The pLV-EF1α-(turboGFP-Luc2)-WPRE transfer vector, the p8.74 packaging vector and the pLV-CMV-(S-ΔRS-HA)-IRES-Puro-WPRE pseudotyping vector were co-transfected in HEK293T cells. Twenty-four and 48 hours post-transfection, Transfected Cells Supernatant (TCS) were harvested, clarified, filtered, aliquoted, stored at −80 °C and subsequently tested on HEK/ACE2/TMPRRS2/Puro cells and on HEK293T cells as negative control. Since pLV-EF1α-(turboGFP-Luc2)-WPRE transfer vector could simultaneously deliver two reporter genes, turboGFP and a human codon usage adapted luciferase (Luc2), cell transduction could be detected either by fluorescence or luminometry. In fact, when different amount of TCS were tested on different numbers of HEK/ACE2/TMPRRS2/Puro cells, an efficient cell transduction could be observed but not on HEK293T negative control cells (Fig. 2A, 2B and C). Further, bonafide fusogenic activity through syncytia formation could be observed (Fig. 2D and E), recapitulating natural CoV-2 infection. Syncytia formation could be considered a signature of CoV infection and pathogenicity since it can be observed not only *in vitro* but also *in vivo*, in lung tissue from people infected with different CoVs, such as SARS-CoV, MERS-CoV or SARS-CoV-2 [19-24].

**Fig. 2.**
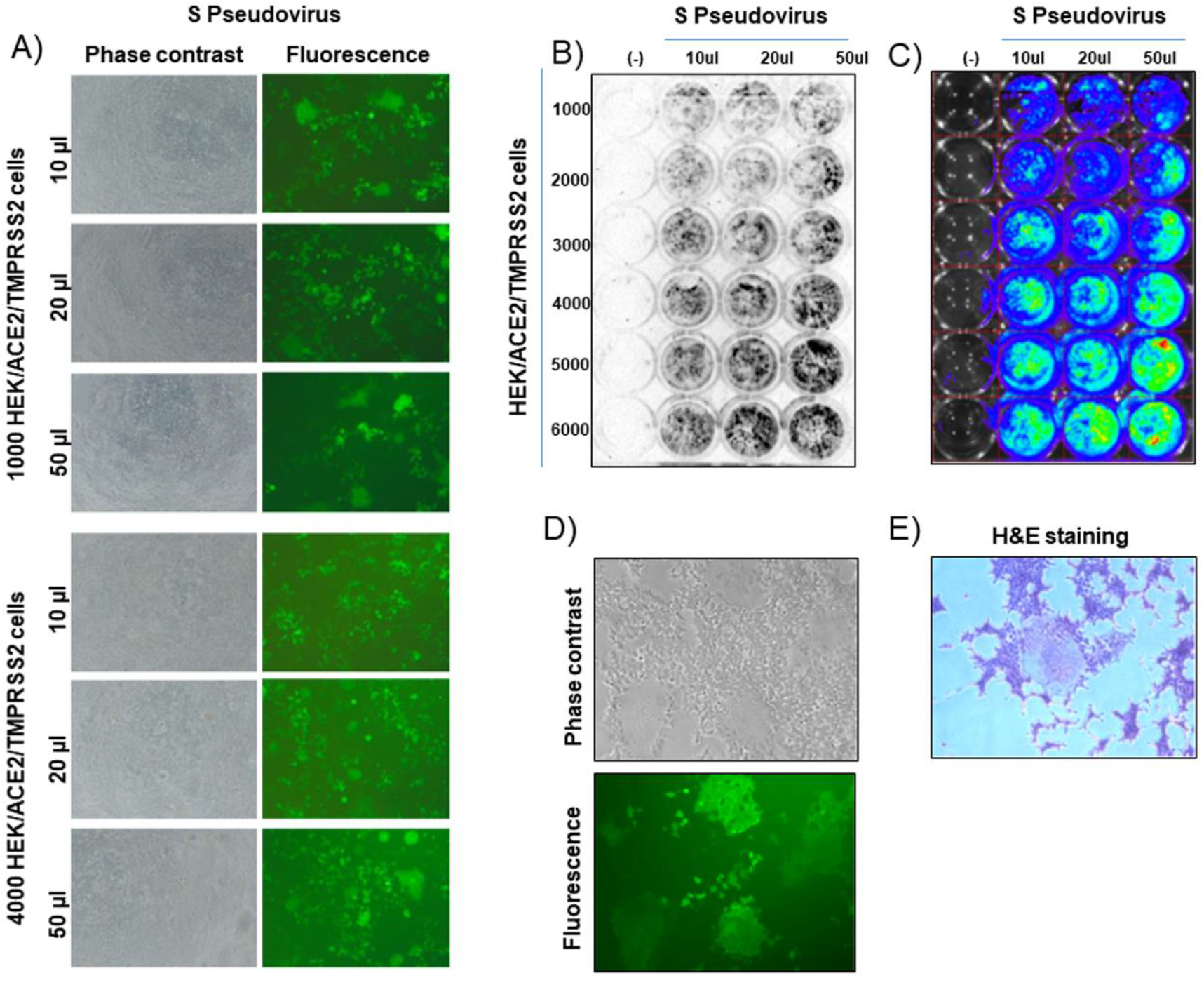
Pseudovirus assembly and transduction. A) Representative images (Phase contrast and fluorescence; 10X) of HEK/ACE2/TMPRRS2/Puro cells, at different number (1000 and 4000), transduced with different amount (50 µL, 20 µL and 10 µL) of supernatant containing pseudovirus. B) ChemiDoc and IVIS (C) luminometric detection of HEK/ACE2/TMPRRS2/Puro cells, at different number (1000, 2000, 3000, 4000, 5000 and 4000), transduced with different amount (50 µL, 20 µL and 10 µL) of supernatant containing pseudovirus (S Pseudovirus). Negative control was established with HEK293T cells, at different number (1000, 2000, 3000, 4000, 5000 and 4000) and transduced with 50 µl of the same supernatant containing pseudovirus (S Pseudovirus). D) Representative images of syncytia (Phase contrast and fluorescence of the same field; 10X), generated by pseudovirus transduced HEK/ACE2/TMPRRS2/Puro cells. E) Representative images of a syncytium (Phase contrast; 10X) generated by pseudovirus transduced HEK/ACE2/TMPRRS2/Puro cells and stained with Hematoxylin and Eosin (H&E staining).

### Lentivirus-based vector can be pseudotyped with ACE2/TMPRSS2

TMPRSS2 is a serine protease with a class II transmembrane domain, whereas ACE2 is a carboxypeptidase with a class I transmembrane domain and both of them are localized on the cell membrane surface [7] (Supplementary Fig. 4). Based on this information it was reasoned that lentiviral vector particles could potentially be pseudotyped with ACE2/TMPRSS2 and allow pseudovirus penetration into cells expressing CoV-2 S. The pLV-EF1α-(turboGFP-Luc2)-WPRE transfer vector, the p8.74 packaging vector and the pEF1α-ACE2/TMPRRS2/Puro pseudotyping vector were co-transfected in HEK293T cells. Twenty-four and 48 hours post transfection TCS were harvested, aliquoted, stored at −80 C° and subsequently tested on pLV-CMV-(S-ΔRS-HA)-IRES-Puro-WPRE transiently or stably transfected HEK/S-ΔRS-HA/Puro cells and on HEK293T cells as negative control. TCS transduction could be observed on HEK/S-ΔRS-HA/Puro cells but not on HEK293T cells negative control cells (Fig. 3A and B). Therefore, a lentivirus-based vector could be pseudotyped with ACE2/TMPRSS2. Worth of note, this kind of pseudotypization and cell transduction did not generate syncytia as observed for the lentivirus-based vector pseudotyped with CoV-2 S. Although the reason of this issue was not investigated, because out of the purpose of this work, it could be assumed that the syncytia formation is an active process in part dependent by a specific intracellular apparatus, contacting the long cytoplasmic tail of ACE2 and/or TMPRSS2, present inside the cells but absent inside the pseudotyped lentiviral particles. It was shown that infection with CoV-2 was significantly decreased in cells expressing ACE2 mutant versions lacking the cytoplasmic domain [25].

**Fig. 3.**
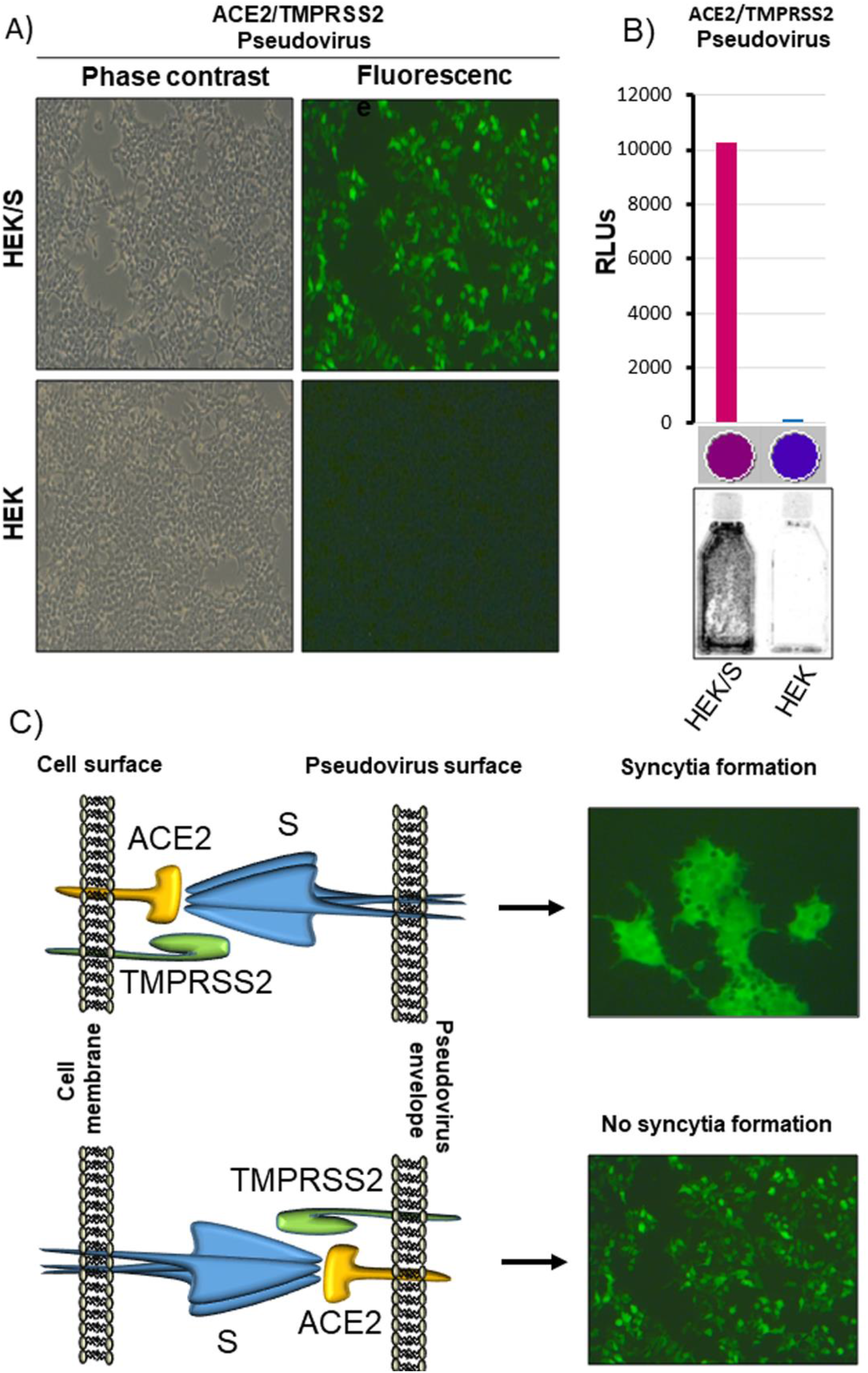
Pseudovirus generation with ACE2 and TMPRSS2. A) Representative image (Phase contrast and Fluorescence of the same field; 10X) of HEK/S-ΔRS-HA/Puro cells (HEK/S) and HEK293T cells (HEK) transduced with ACE2/TMPRSS2 Pseudovirus. B) Representative 25 cm2 flasks of HEK/S-ΔRS-HA/Puro cells (HEK/S) and HEK293T cells (HEK) transduced with ACE2/TMPRSS2 Pseudovirus and exposed to the ChemiDoc apparatus. Luminometric quantification of the lysate of the same cells, expressed as Relative Luciferase Units (RLUs) and colorimetrically visualized (violet and blue spots) by the luminometer software, during and after reading. C) Cartoon explaining the lack of syncytia generation by ACE2/TMPRSS2 Pseudovirus.

### Assessment of a pseudovirus neutralization assay

Starting from the availability of a pseudovirus mimicking the interaction of SARS-CoV-2 with target cells, it was of interest to assess a pseudovirus neutralization assay, to be applied for several purposes. The aim of this assay is to overcome the other neutralization assays complexities, such as multiple passages and the addition and removal of reagents and solutions, mainly during sera dilution and detection steps, where cells could detach from the microplate surface and strong errors could be introduced. In our simplified system reagents and solution are only added at each step, while washing steps are absent, from the beginning to the end of the assay.

Opaque wall, clear flat bottom 96 well microplates were employed (Supplementary Fig. 5). Opaque walls prevent well-to-well crosstalk for luminometric detection whereas clear flat bottom well direct microscopic monitoring cell growth and/or fluorescence microscope observation of transduced cells expressing a fluorescent reporter gene, in this specific case turboGFP. The neutralization assay was performed with human sera coming from SARS-CoV-2-infected patients, collected after post–symptoms onset, that resulted positive to PCR and ELISA tests. Whereas, sera collected prior to the SARS-CoV-2 emergence (1999) that tested negative in ELISA were used as negative control. First, serial dilution of the sera (plasma from EDTA treated blood can be used too), were incubated for 90 minutes with TCS containing pseudovirus sufficient to achieve approximately 10^4^ Relative Light Units (RLUs) of luciferase signal per well (usually no more than 2 to 10 µL). Subsequently 10^4^ HEK/ACE2/TMPRRS2/Puro cells were added to each well (Supplementary Fig. 5). As previously mentioned, HEK/ACE2/TMPRRS2/Puro cells do not need to be constantly kept in culture under puromycin selection: a large batch of them can be grown, aliquoted at a suitable concentration, stored at −80 C° or in liquid nitrogen and be immediately ready to use after thawing. Alternatively, HEK293T cells can be electroporated with pEF1α-ACE2/TMPRRS2/Puro and directly used, obtaining the same result. NT50 could be measured as soon as 48 hours after adding the cells, either by a fluorescence microscope, or by luminometry and simply adding Luciferine in each well (Supplementary Fig. 5 and Fig. 4A and B), without the need of cell lysis and of transferring the lysate to a different microplate. Moreover, light detection can be performed with a Digital Imager for western blotting, BLI, or by a multiwell plate reader luminometer.

**Fig. 4.**
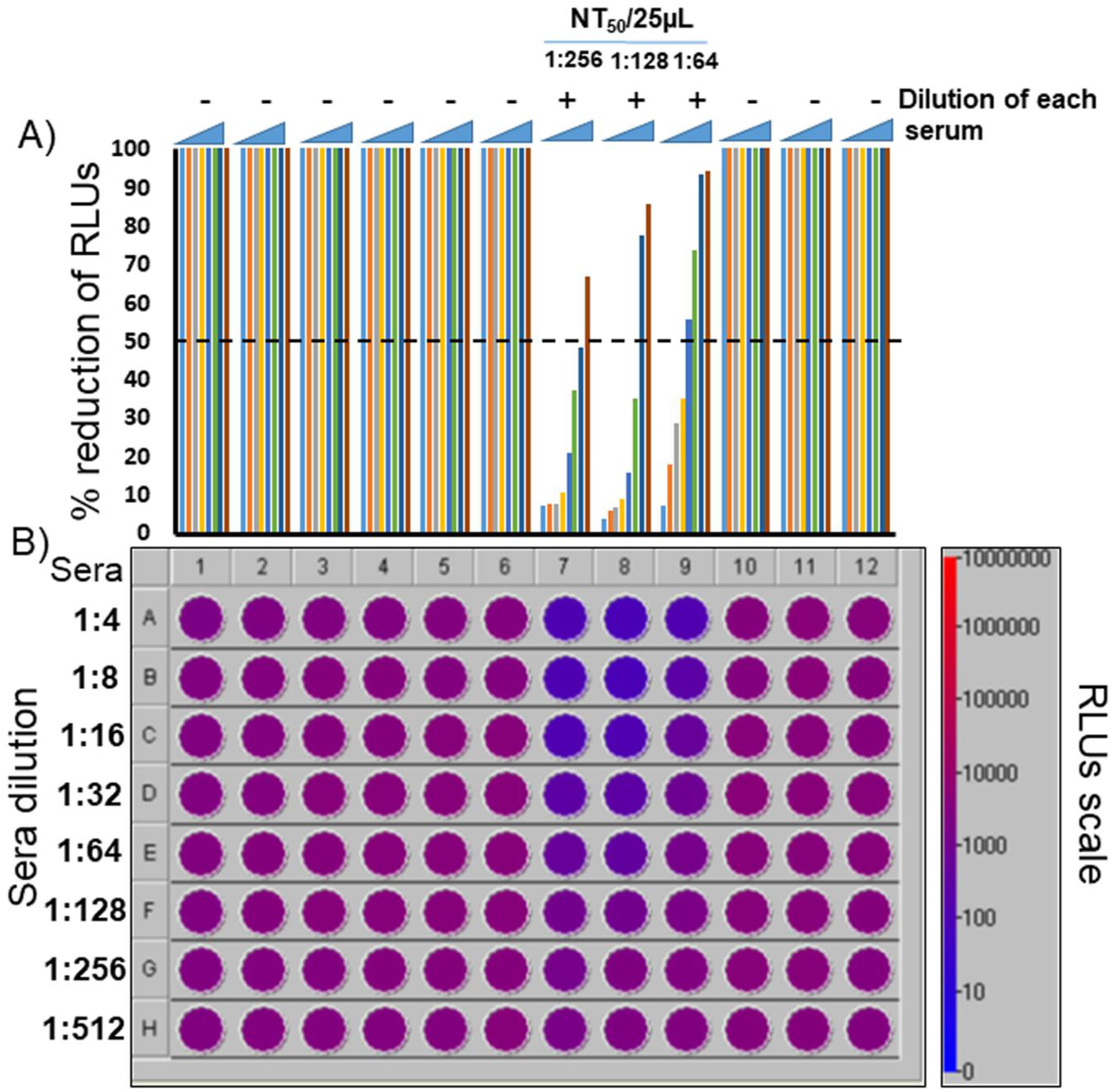
Representative pseudovirus neutralization assay. Six PCR and ELISA CoV2 negative patients’ representative sera (1 to 6), 3 PCR and ELISA CoV2 positive patients’ sera (7 to 9) and 2 pre-pandemic sera (collected in the 1998; 10 and 11) were tested. A further negative control was established without serum (12), which was substituted with complete medium. (A) Histogram obtained from luminometric detection of transduced cells with pseudovirus. Relative Luciferase Units (RLUs) were compared and normalized to those derived from wells where pseudovirus were added in the absence of sera (100%). Neutralization titer 50 (NT_50_) was expressed as the maximal dilution of the sera where the reduction of the signal was ≥50%. In this specific example, the NT_50_/25µL of the serum 7 is 1:256, that of the serum 8 is 1:128 and that of the serum 9 is 1:64. However, because the titer was measured on 25 µL, the apparent NT_50_ has to be multiplied per 40 to get the normalized NT_50_/mL. Therefore, the NT_50_/mL of the sera 7 is 1:10256, that of the serum 8 is 1:5120 and that of the serum 9 is 1:2560. (B) Colorimetric visualization of the RLUs displayed by the luminometer software, during and after reading.

Thus, a rapid, flexible, reliable, sensitive and cost effective pseudovirus neutralization assay for SARS-CoV-2 was established, where all steps could be performed in the same plate and in a standard BSL2 laboratory, since the pseudovirus is completely safe. With this assay we have been able to rapidly and cheaply determine the neutralizing potency of mAbs derived from serum or plasma samples in a BSL2 laboratory. Automation and additional miniaturization is certainly possible to further increase throughput. Since serum neutralizing activity has long been identified as a correlate of protection in many viral infection [26], including CoVs [27] and a global CoV-2 vaccination campaign is ongoing, it is absolutely important to have a practical test to assess the outcome of vaccination. In order to meet this need, such test must be rapid, flexible, reliable, sensitive, quantitative, cost effective, safe and easily adapted to different coronaviruses variants.

## Data Availability

All data are available

## Acknowledgement

We would like to thank all the people who decided to donate their blood for the study and Diletta Laccabue, Laboratory of Viral Immunopathology, Unit of Infectious Diseases and Hepatology, Azienda Ospedaliero-Universitaria di Parma, for taking care of the experimental protocol submission to the ethical committee.

---

**Supplementary Figure 1.**
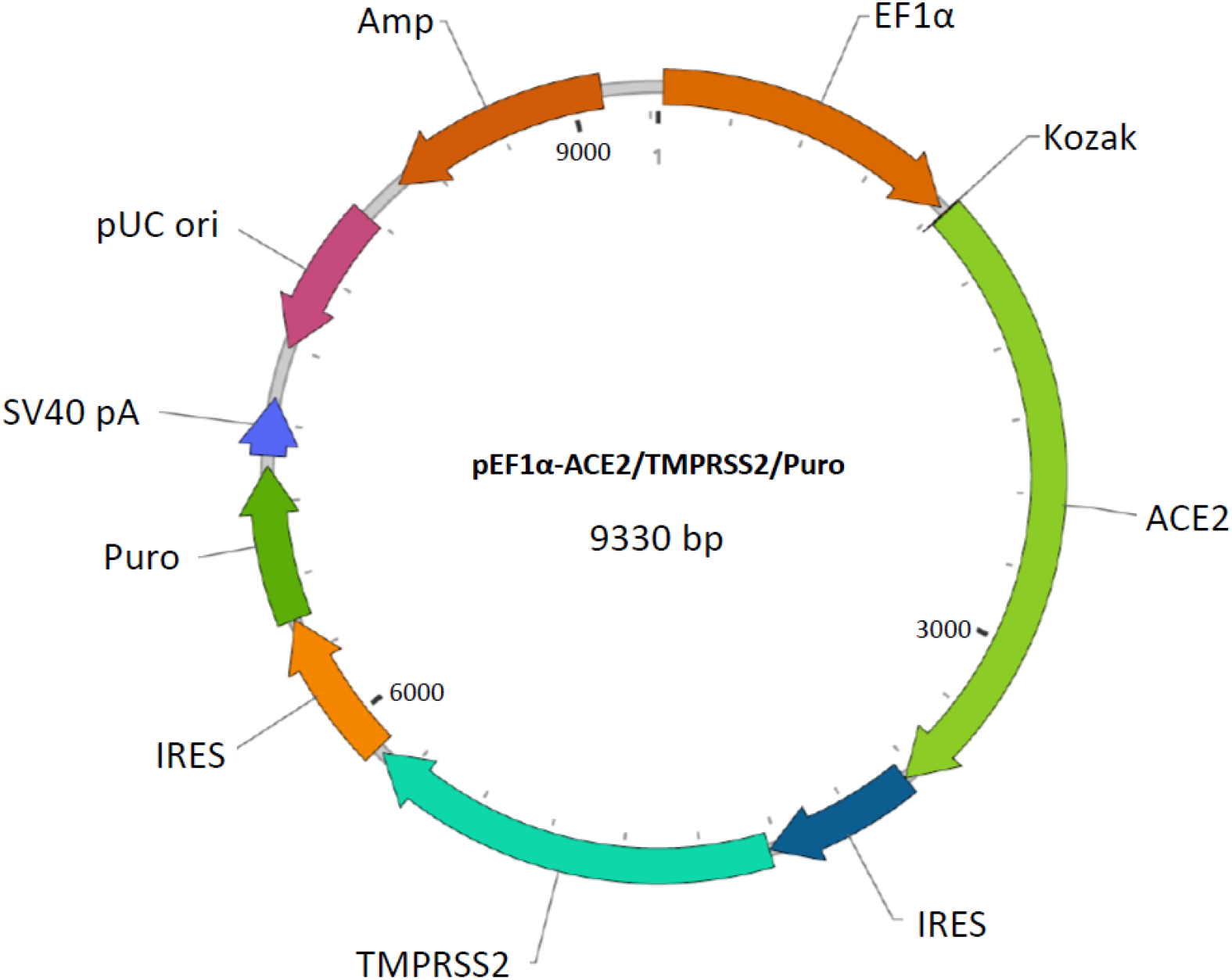
Diagram not on scale of pEF1α-ACE2/TMPRSS2/Puro construct, where functional elements of the construct are indicated and highlighted with different colours. EF1α (human Elongation Factor 1 alpha promoter), Kozak (Kozak’s translation initiation sequence), ACE2 (Human Angiotensin Converting Enzyme 2 ORF), IRES (Encephalomyocarditis virus internal ribosomal entry site), TMPRSS2 (human Transmembrane Protease Serine 2), Puro (Puromycin resistance gene ORF), pUC ori (High copy number pUC origin of replication) SV40 pA (Simian virus 40 early polyadenylation signal) and Amp (Ampicillin resistance gene, comprising the prokaryotic promoter and ORF).

**Supplementary Figure 2.**
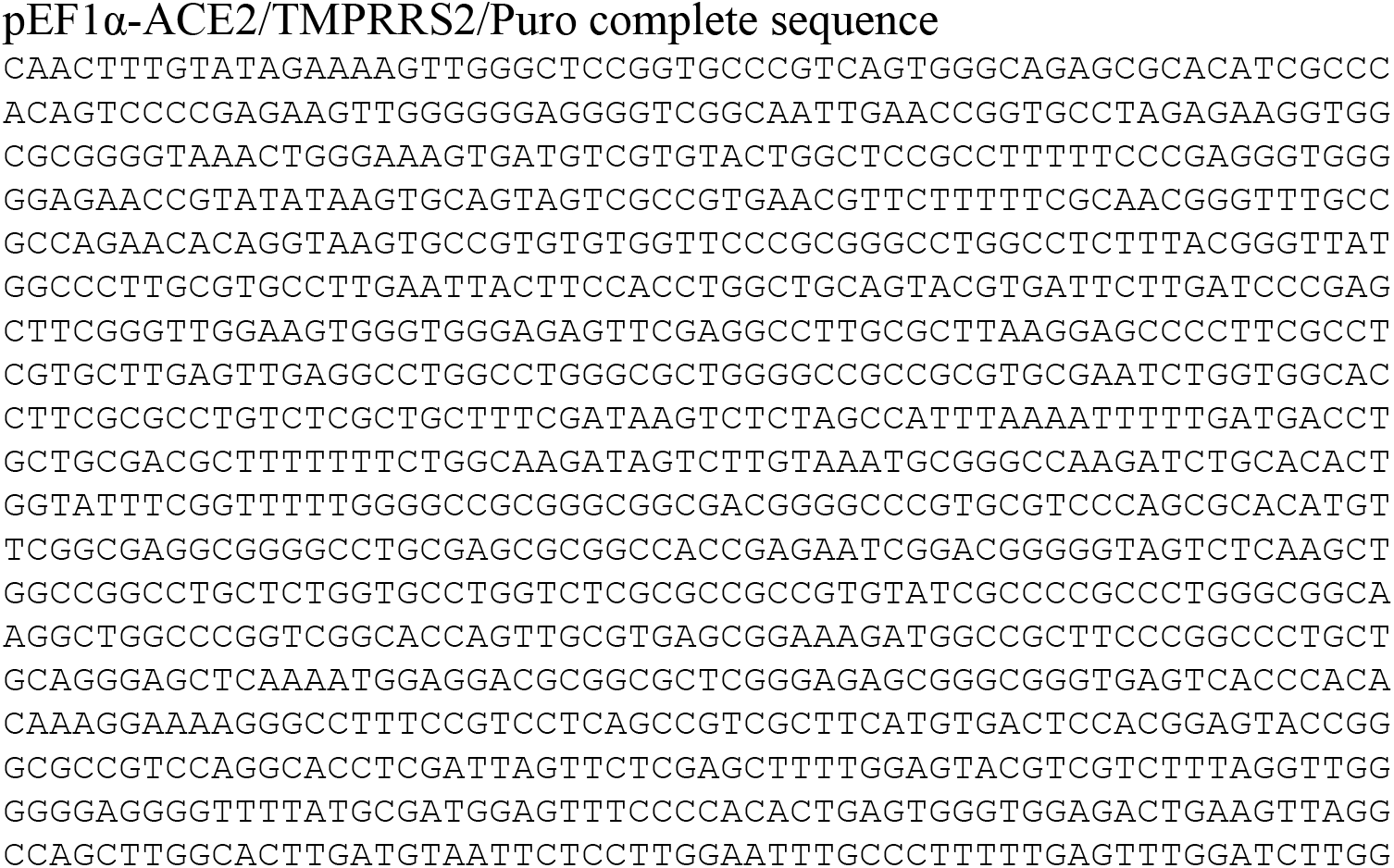
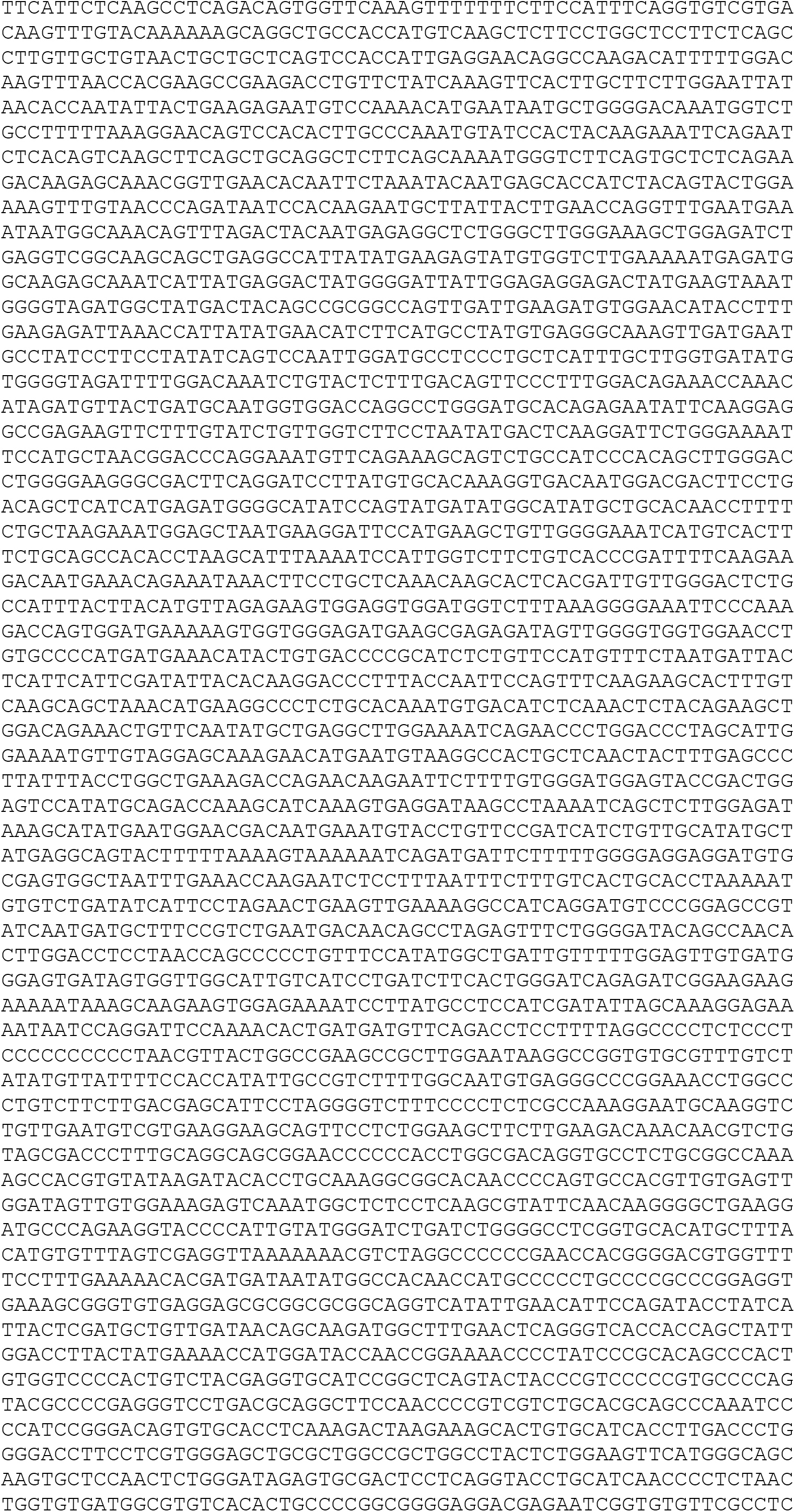
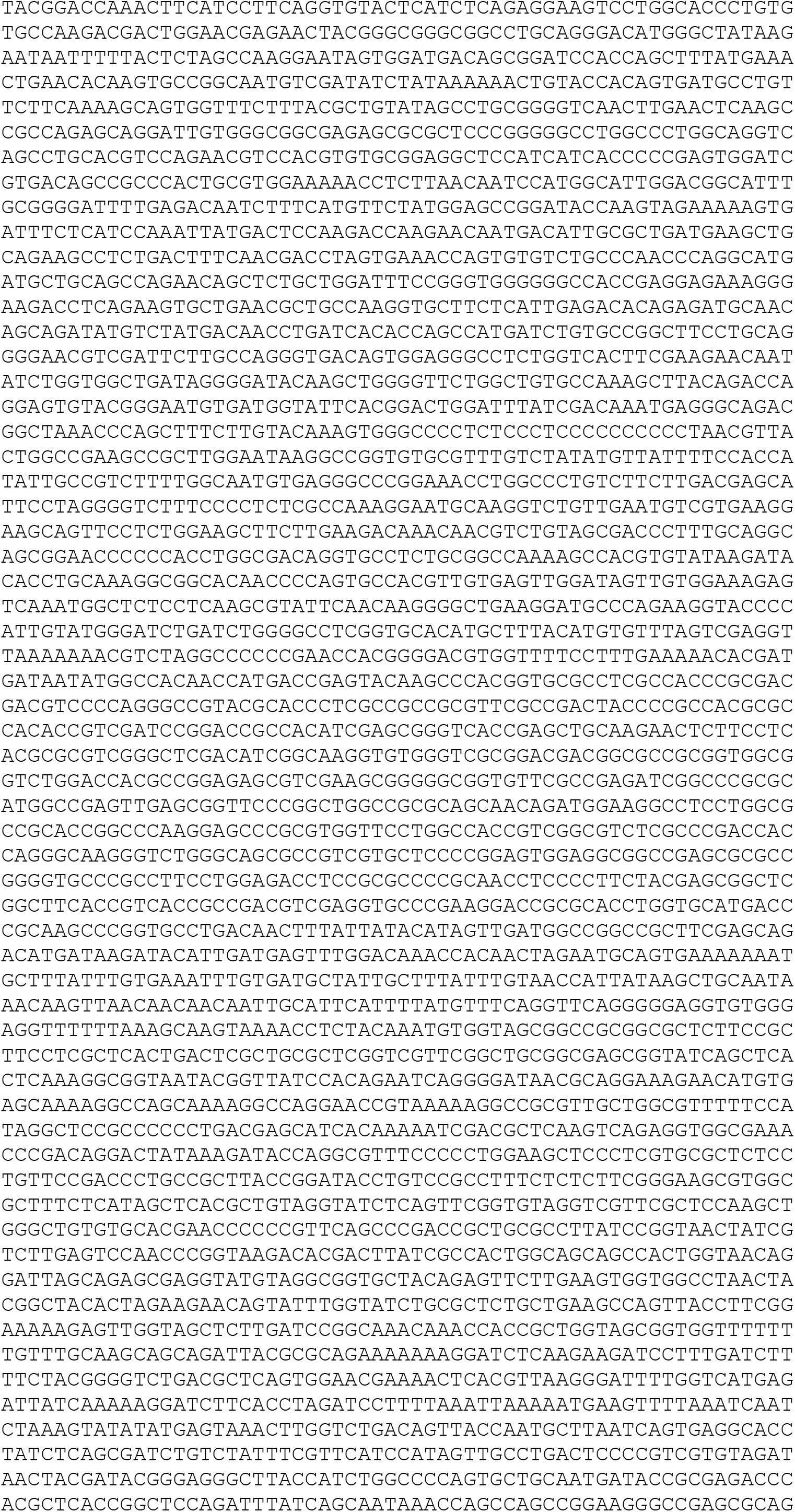
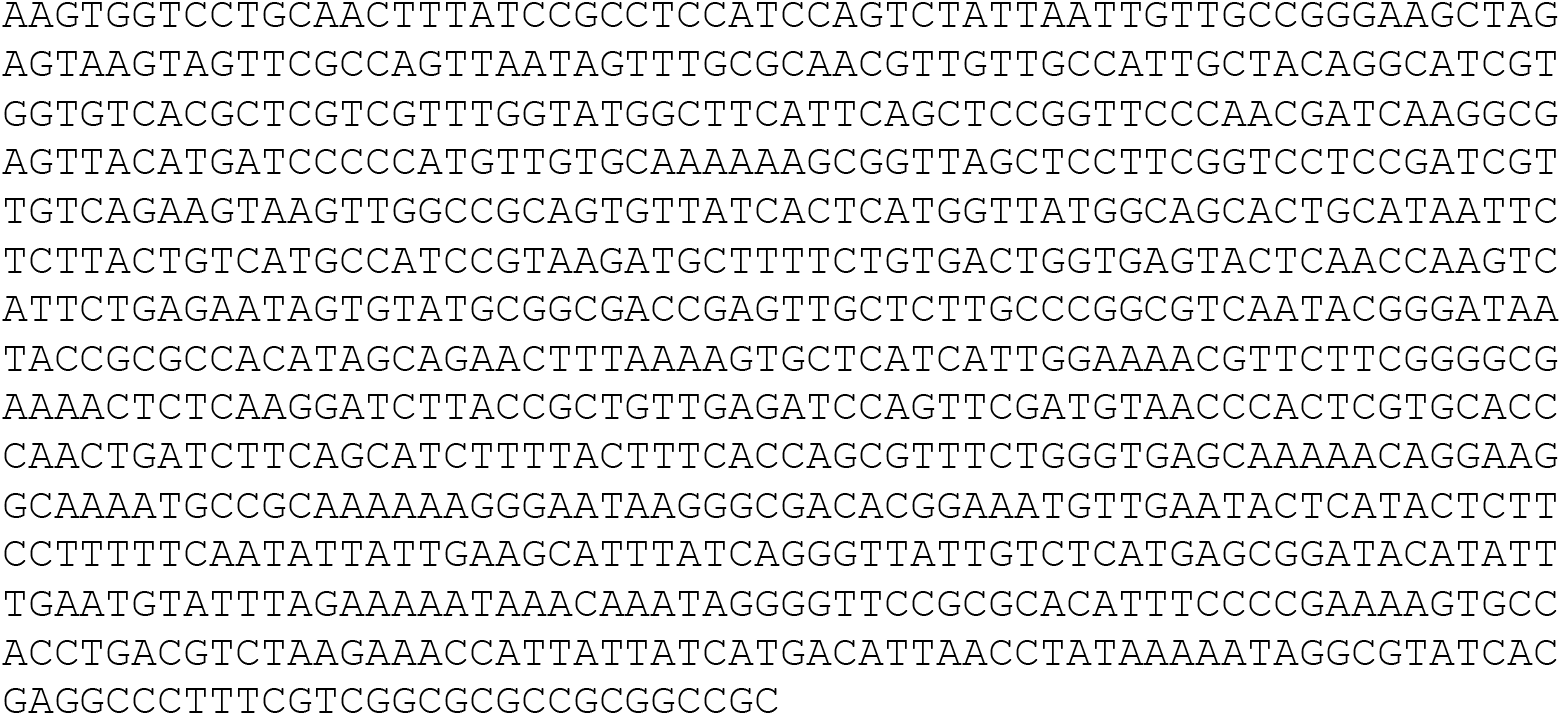

**Supplementary Figure 3.**
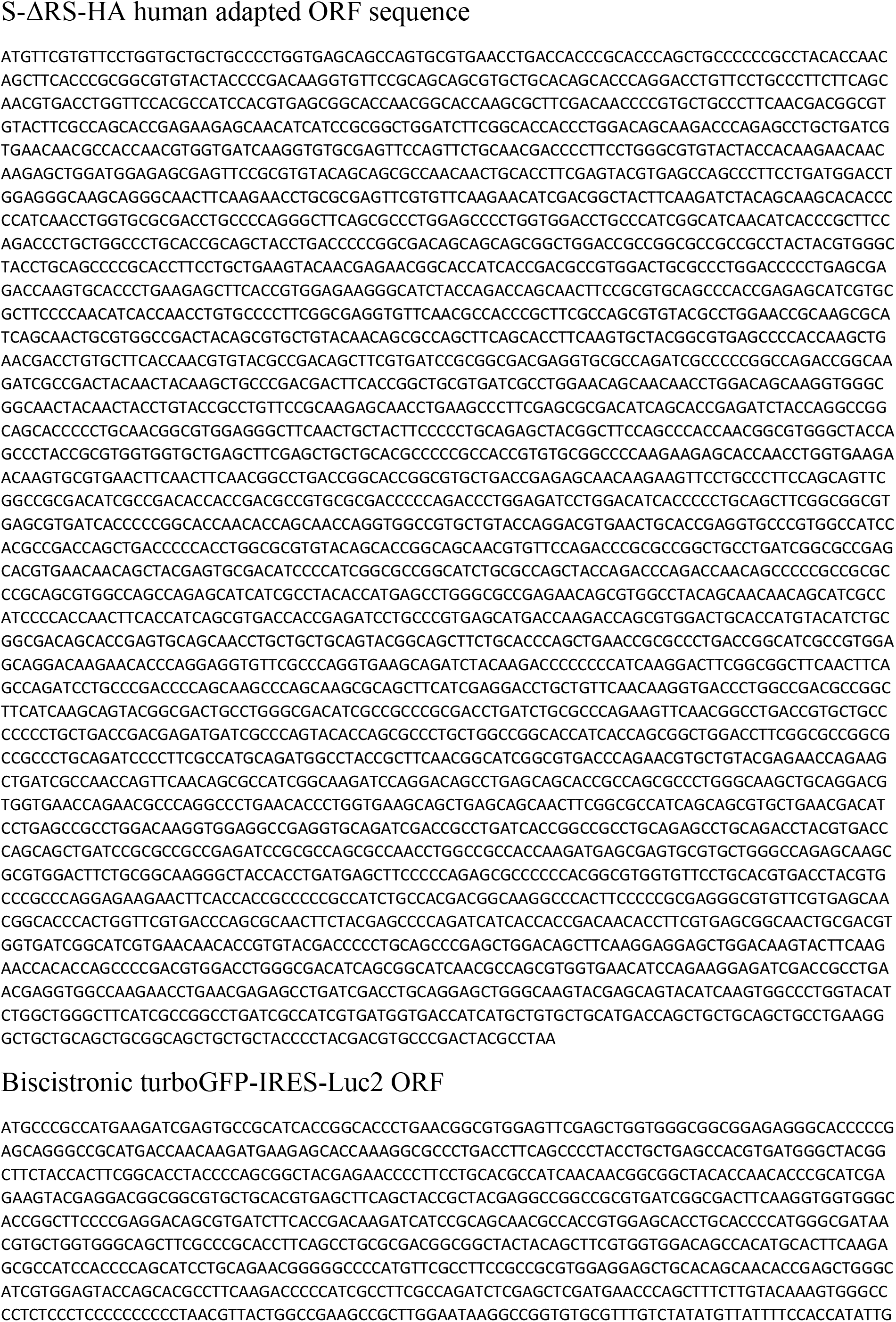
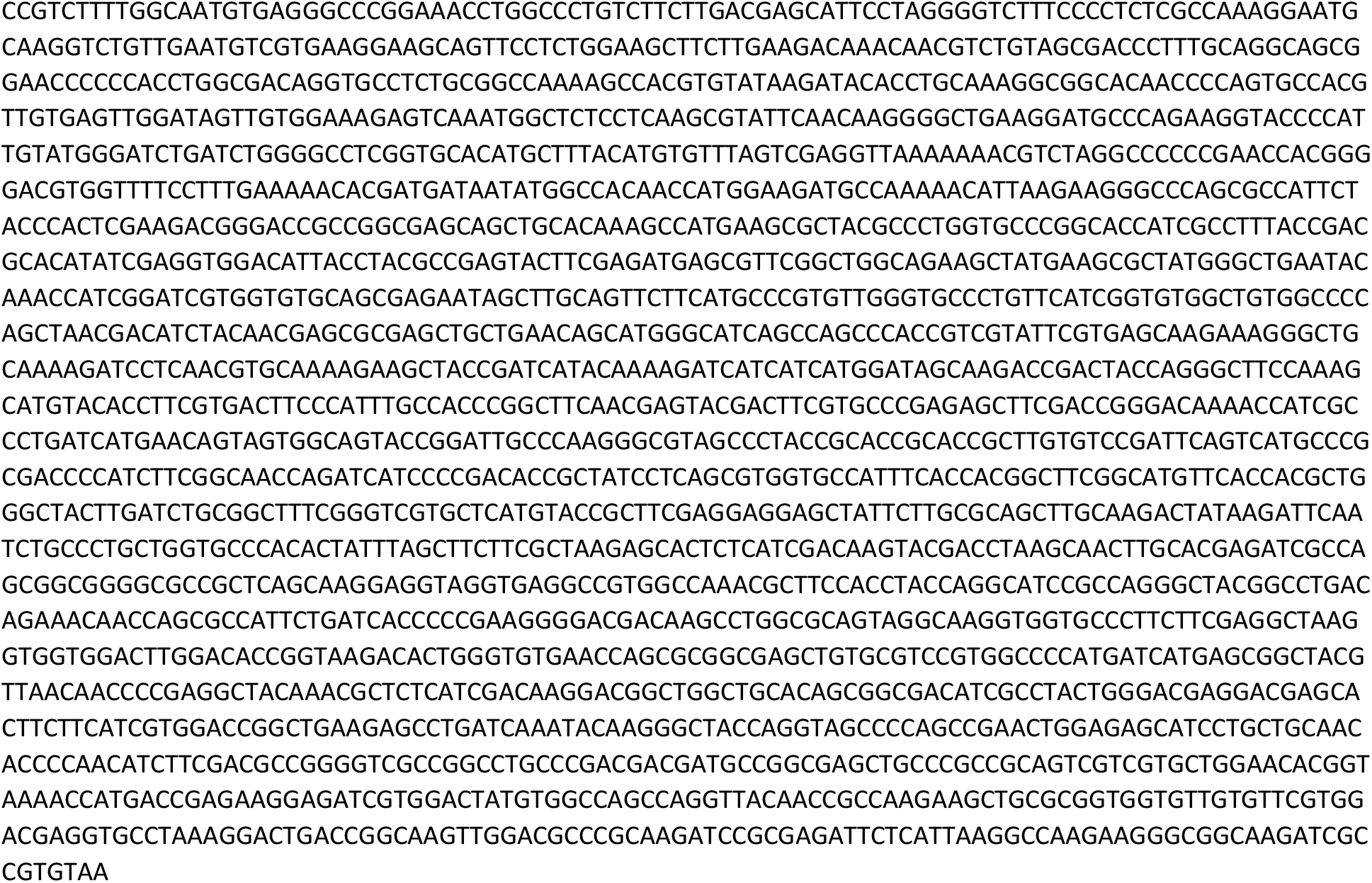

**Supplementary Figure 4.**
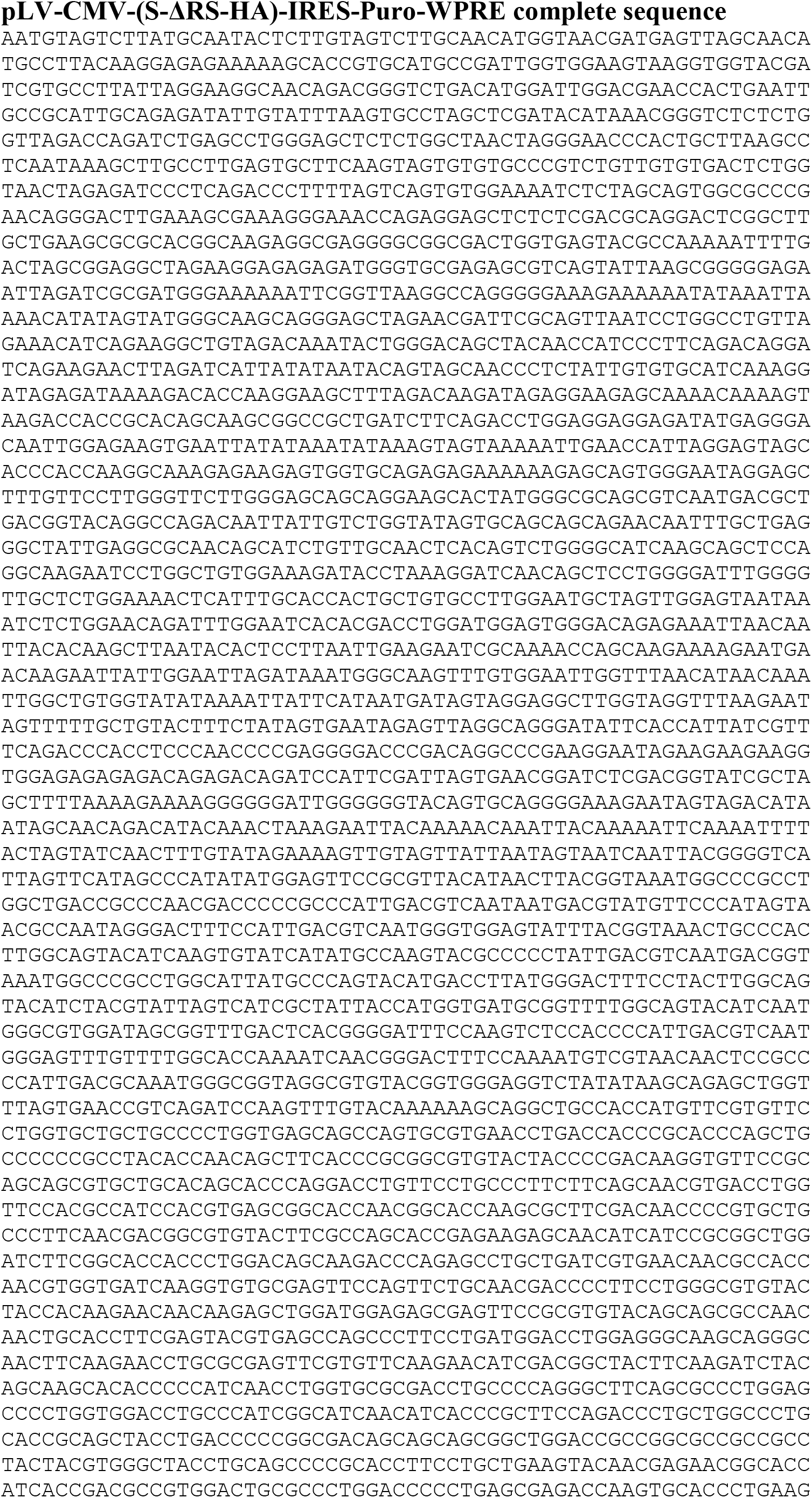
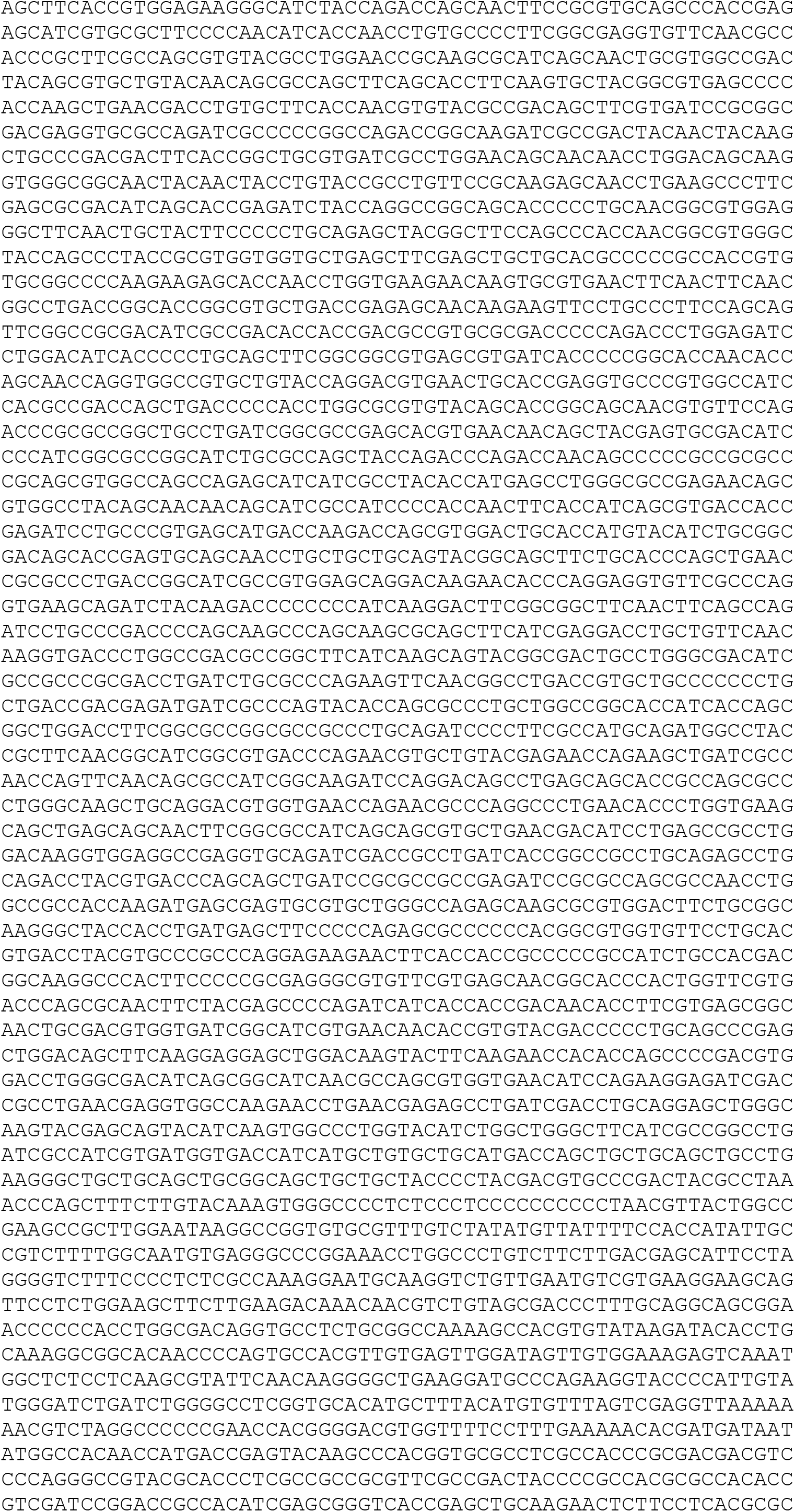
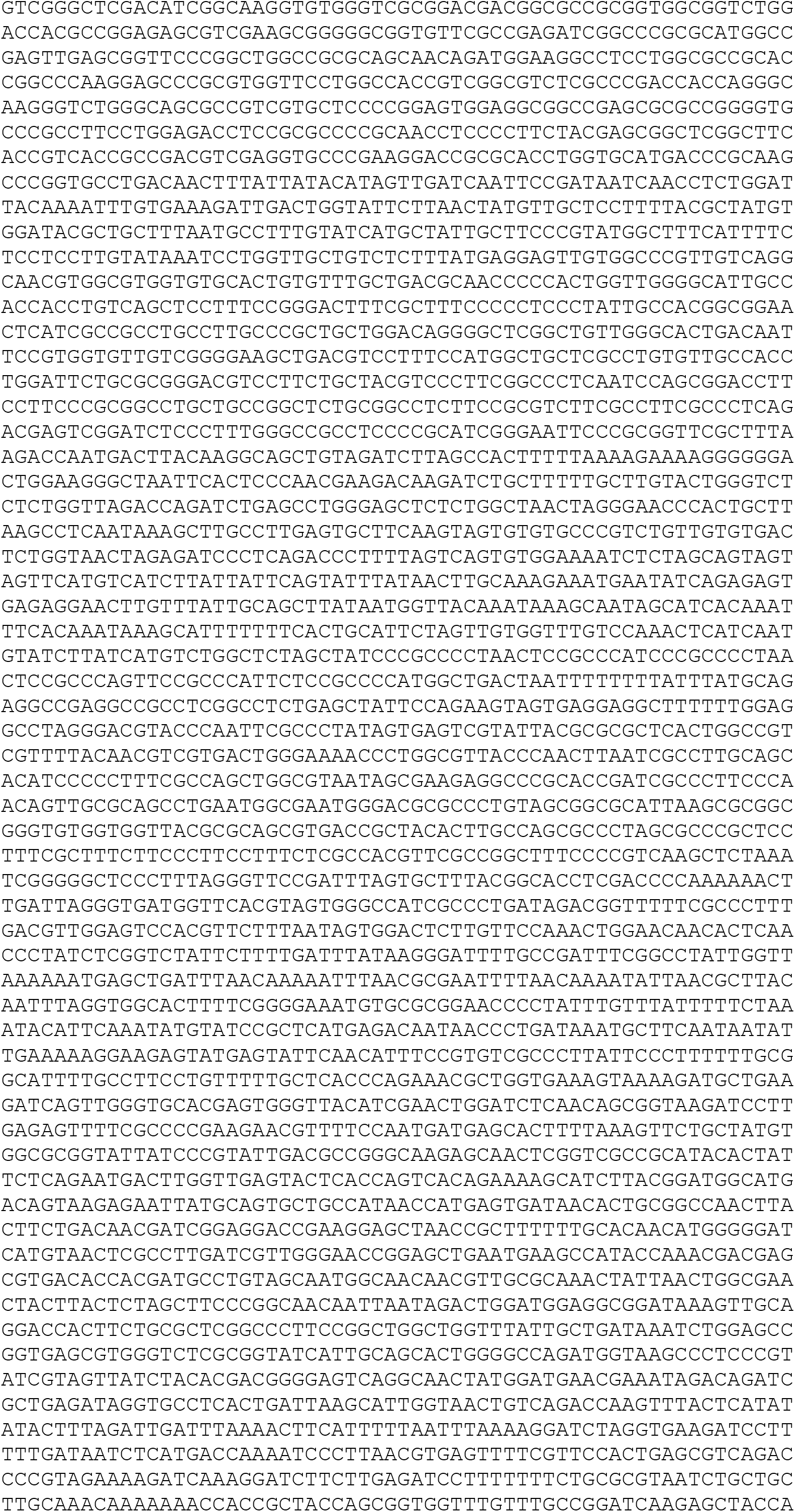
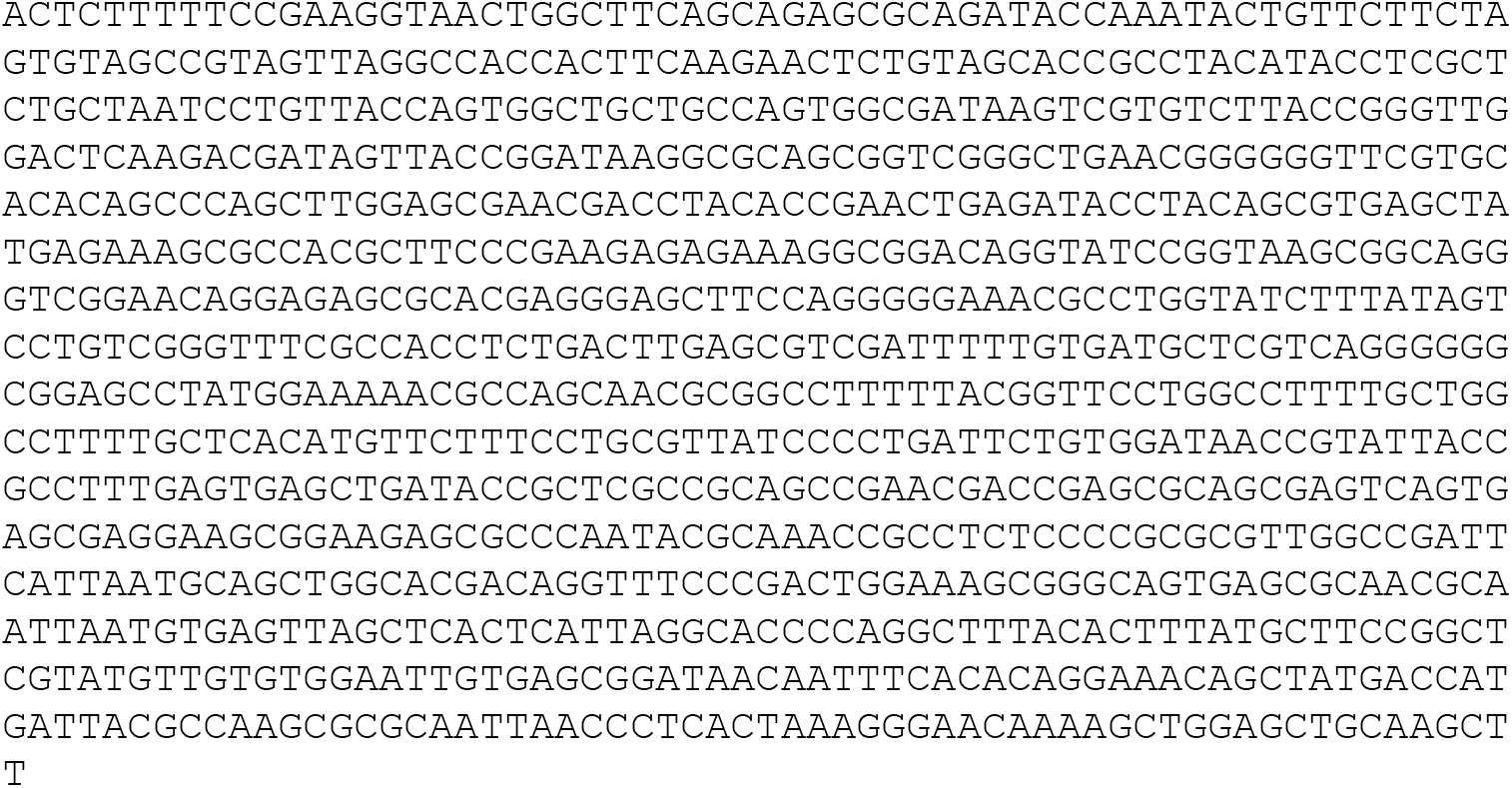
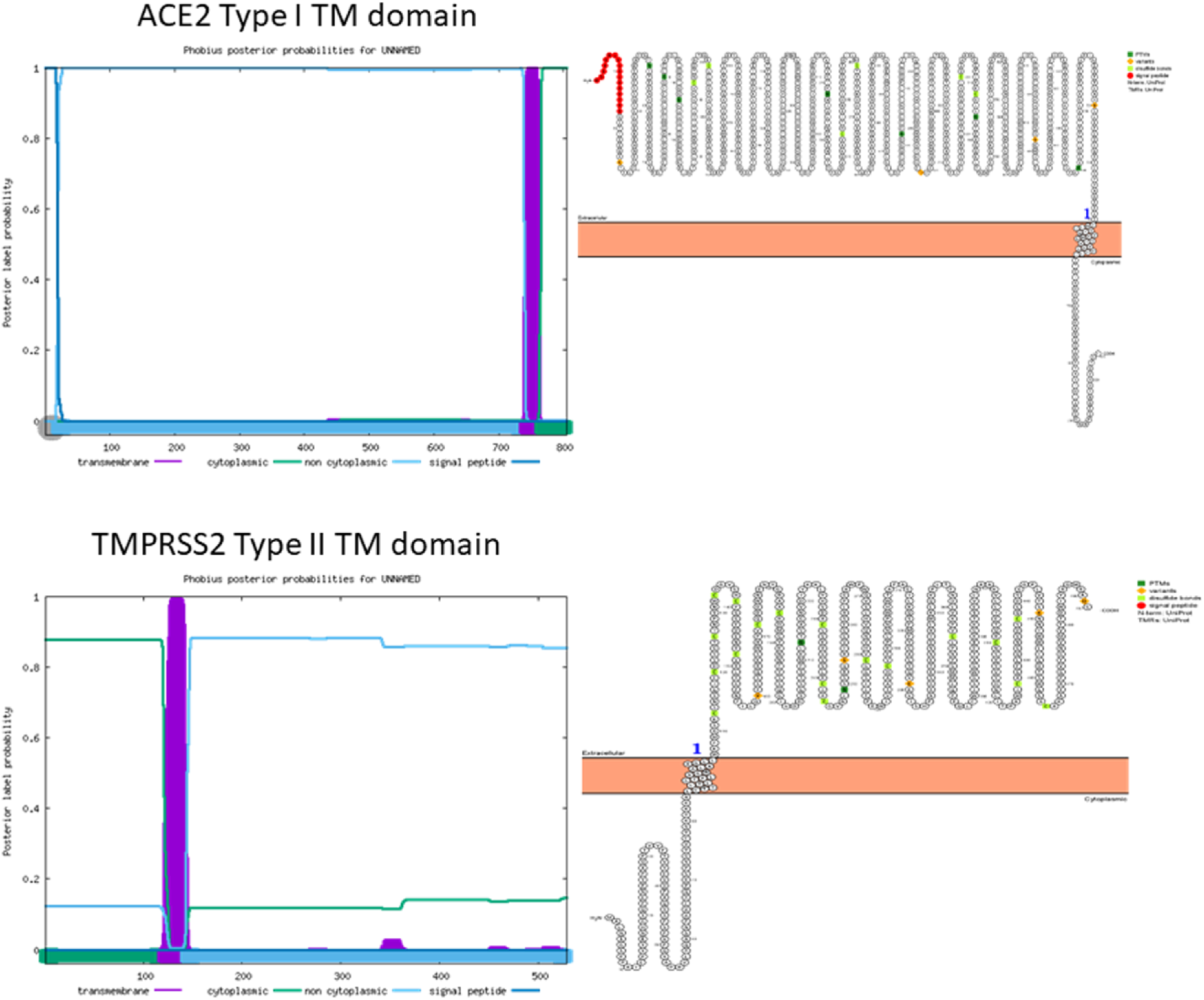
A) A combined transmembrane topology and signal peptide prediction of ACE2 and TMPRSS2 as performed by Phobius (https://phobius.sbc.su.se). B) A topological and subcellular localization prediction of ACE2 and TMPRSS2 as performed by Protter (https://wlab.ethz.ch/protter/start/). Open-source tool for visualization of proteoforms and interactive integration of annotated and predicted sequence features together with experimental proteomic evidence.

**Supplementary Figure 5.**
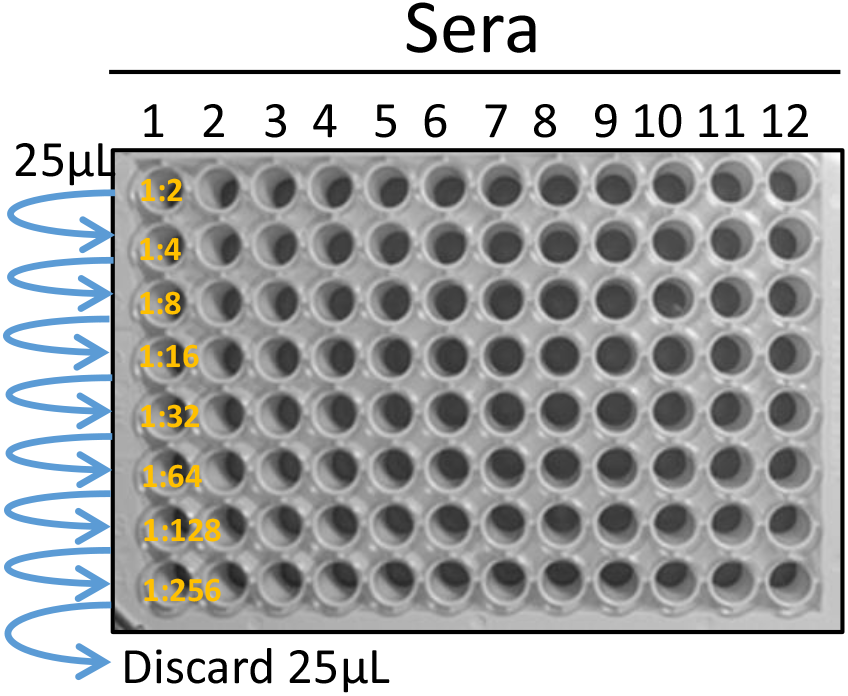
1) Dilution of the sera 25µL of complete medium are added to each well of the microplate and 25 µL of each serum is added to each well of the first line of wells. Than 25 µL of the first dilution of the sera (1:2) are mixed and passed to the subsequent lines of wells. 25µL from the last line of wells are discarded. Therefore, the final volume for each well is 25µL. 2) Adding the pseudovirus 25µL of pseudovirus in complete medium (corresponding to 10^4^ RLUs; ∼3-5µL of the initial preparation) are added to each well and left at room temperature for 1,5 hours. Final volume for each well is 50µL, therefore the sera dilution is doubled (1:4-1:8-1:16-1:32-1:64-1:128-1:256-1:512). 3) Adding the cells 50µL of complete medium containing 10^4^ HEK/ACE2/TMPRRS2/Puro cells are added to each well and left for 1,5 hours in cell incubator. Final volume for each well is 100µL. 4) Adding the Luciferine and reading 25µL of complete medium containing Luciferine (20µL of complete medium plus 5µL of 15mg/mL Luciferine in PBS) are added to each well just before the luminometric reading of the microplate. Final volume for each well is 125µL. Since the plate has a transparent bottom, a dark film layer should be applied for reading. RLUs obtained were compared and normalized to those derived from wells where pseudovirus were added in the absence of plasma/sera (100%). Neutralization titer 50 (NT_50_/mL) was expressed as the maximal dilution of the sera where the reduction of the signal is ≥50%. This titer has to be multiplied per 40 because the initial volume of the sera tested is 0,025mL and it has to be normalized to 1mL. This is a very important issue because SN protocol could differ from each lab and the initial amount of sera tested could be different in volume. However, in most of the case, operators tend to indicate SN_50_ and not SN_50_/mL.

## References

1. Han Q, Lin Q, Jin S, et al. Coronavirus 2019-nCoV: A brief perspective from the front line. J Infect. 2020 Apr;80(4):373–377.

2. Callaway E. The race for coronavirus vaccines: a graphical guide. Nature. 2020 Apr;580(7805):576–577.

3. Poland GA, Ovsyannikova IG, Kennedy RB. SARS-CoV-2 immunity: review and applications to phase 3 vaccine candidates. Lancet. 2020 Nov 14;396(10262):1595–1606.

4. Iacob S, Iacob DG. SARS-CoV-2 Treatment Approaches: Numerous Options, No Certainty for a Versatile Virus. Front Pharmacol. 2020;11:1224.

5. Li G, De Clercq E. Therapeutic options for the 2019 novel coronavirus (2019-nCoV). Nat Rev Drug Discov. 2020 Mar;19(3):149–150.

6. Walls AC, Park YJ, Tortorici MA, et al. Structure, Function, and Antigenicity of the SARS- CoV-2 Spike Glycoprotein. Cell. 2020 Apr 16;181(2):281–292 e6.

7. Hoffmann M, Kleine-Weber H, Schroeder S, et al. SARS-CoV-2 Cell Entry Depends on ACE2 and TMPRSS2 and Is Blocked by a Clinically Proven Protease Inhibitor. Cell. 2020 Apr 16;181(2):271–280 e8.

8. Yan R, Zhang Y, Li Y, et al. Structural basis for the recognition of SARS-CoV-2 by full-length human ACE2. Science. 2020 Mar 27;367(6485):1444–1448.

9. Lin K, Liu M, Ma H, et al. Laboratory biosafety emergency management for SARS-CoV-2. J Biosaf Biosecur. 2020 Sep 19.

10. Case JB, Rothlauf PW, Chen RE, et al. Neutralizing Antibody and Soluble ACE2 Inhibition of a Replication-Competent VSV-SARS-CoV-2 and a Clinical Isolate of SARS-CoV-2. Cell Host Microbe. 2020 Sep 9;28(3):475–485 e5.

11. Nie J, Li Q, Wu J, et al. Establishment and validation of a pseudovirus neutralization assay for SARS-CoV-2. Emerg Microbes Infect. 2020 Dec;9(1):680–686.

12. Crawford KHD, Eguia R, Dingens AS, et al. Protocol and Reagents for Pseudotyping Lentiviral Particles with SARS-CoV-2 Spike Protein for Neutralization Assays. Viruses. 2020 May 6;12(5).

13. Giroglou T, Cinatl J, Jr., Rabenau H, et al. Retroviral vectors pseudotyped with severe acute respiratory syndrome coronavirus S protein. J Virol. 2004 Sep;78(17):9007–15.

14. Ujike M, Huang C, Shirato K, et al. The contribution of the cytoplasmic retrieval signal of severe acute respiratory syndrome coronavirus to intracellular accumulation of S proteins and incorporation of S protein into virus-like particles. J Gen Virol. 2016 Aug;97(8):1853–1864.

15. Jouvenet N, Neil SJD, Bess C, et al. Plasma membrane is the site of productive HIV-1 particle assembly. Plos Biology. 2006 Dec;4(12):2296–2310.

16. Gustafsson C, Govindarajan S, Minshull J. Codon bias and heterologous protein expression. Trends Biotechnol. 2004 Jul;22(7):346–53.

17. Kudla G, Lipinski L, Caffin F, et al. High guanine and cytosine content increases mRNA levels in mammalian cells. PLoS Biol. 2006 Jun;4(6):e180.

18. Ziegler CGK, Allon SJ, Nyquist SK, et al. SARS-CoV-2 Receptor ACE2 Is an Interferon- Stimulated Gene in Human Airway Epithelial Cells and Is Detected in Specific Cell Subsets across Tissues. Cell. 2020 May 28;181(5):1016–1035 e19.

19. Buchrieser J, Dufloo J, Hubert M, et al. Syncytia formation by SARS-CoV-2-infected cells. EMBO J. 2020 Oct 13:e106267.

20. Bussani R, Schneider E, Zentilin L, et al. Persistence of viral RNA, pneumocyte syncytia and thrombosis are hallmarks of advanced COVID-19 pathology. EBioMedicine. 2020 Nov;61:103104.

21. Franks TJ, Chong PY, Chui P, et al. Lung pathology of severe acute respiratory syndrome (SARS): a study of 8 autopsy cases from Singapore. Hum Pathol. 2003 Aug;34(8):743–8.

22. Xia S, Liu M, Wang C, et al. Inhibition of SARS-CoV-2 (previously 2019-nCoV) infection by a highly potent pan-coronavirus fusion inhibitor targeting its spike protein that harbors a high capacity to mediate membrane fusion. Cell Res. 2020 Apr;30(4):343–355.

23. Matsuyama S, Nagata N, Shirato K, et al. Efficient activation of the severe acute respiratory syndrome coronavirus spike protein by the transmembrane protease TMPRSS2. J Virol. 2010 Dec;84(24):12658–64.

24. Qian Z, Dominguez SR, Holmes KV. Role of the spike glycoprotein of human Middle East respiratory syndrome coronavirus (MERS-CoV) in virus entry and syncytia formation. PLoS One. 2013;8(10):e76469.

25. Haga S, Yamamoto N, Nakai-Murakami C, et al. Modulation of TNF-alpha-converting enzyme by the spike protein of SARS-CoV and ACE2 induces TNF-alpha production and facilitates viral entry. Proc Natl Acad Sci U S A. 2008 Jun 3;105(22):7809–14.

26. Plotkin SA. Correlates of Protection Induced by Vaccination. Clinical and Vaccine Immunology. 2010 Jul;17(7):1055–1065.

27. Kellam P, Barclay W. The dynamics of humoral immune responses following SARS-CoV-2 infection and the potential for reinfection. Journal of General Virology. 2020;101(8):791–797.

